# Pharmacological inhibition of the kinin-kallikrein system in severe COVID-19 – A proof-of-concept study

**DOI:** 10.1101/2020.08.11.20167353

**Authors:** Eli Mansour, Andre C. Palma, Raisa G. Ulaf, Luciana C. Ribeiro, Ana Flavia Bernardes, Thyago A. Nunes, Marcus V. Agrela, Bruna Bombassaro, Milena Monfort-Pires, Rafael L. Camargo, Eliana P. Araujo, Natalia S. Brunetti, Alessandro S. Farias, Antônio Luís E. Falcão, Thiago Martins Santos, Plinio Trabasso, Rachel P. Dertkigil, Sergio S. Dertkigil, Maria Luiza Moretti, Licio A. Velloso

**Author notes:** Corresponding author; Licio A. Velloso.

## Abstract

Coronavirus disease-19 (COVID-19) can develop into a severe respiratory syndrome that results in up to 40% mortality. Acute lung inflammatory edema is a major pathological finding in autopsies explaining O_2_ diffusion failure and hypoxemia. Only dexamethasone has been shown to reduce mortality in severe cases, further supporting a role for inflammation in disease severity. SARS-CoV-2 enters cells employing angiotensin converting enzyme 2 (ACE2) as a receptor, which is highly expressed in lung alveolar cells. ACE2 is one of the components of the cellular machinery that inactivates the potent inflammatory agent bradykinin, and SARS-CoV-2 infection could interfere with the catalytic activity of ACE2, leading to accumulation of bradykinin. In this open-label, randomized clinical trial, we tested two pharmacological inhibitors of the kinin-kallikrein system that are currently approved for the treatment of hereditary angioedema, icatibant and inhibitor of C1 esterase/kallikrein, in a group of 30 patients with severe COVID-19. Neither icatibant nor inhibitor of C1 esterase/kallikrein resulted in significant changes in disease mortality and time to clinical improvement. However, both compounds promoted significant improvement of lung computed tomography scores and increased blood eosinophils, which has been reported as an indicator of disease recovery. In this small cohort, we found evidence for a beneficial role of pharmacological inhibition of the kinin-kallikrein system in two markers that indicate improved disease recovery.

## Introduction

Acute respiratory failure is the leading cause of death in coronavirus disease-19 (COVID-19) ^1,2^. In severe cases, there is progressive shortening of breath and hypoxemia that often require supplementary oxygen or mechanical ventilation ^2,3^. Chest computed tomography (CT) scans reveal multiple lung opacities that are both central and peripheral and can present as ground-glass and consolidation types, usually involving two or more lobes ^4,5^. Histological examination of lung autopsy specimens identified angiocentric inflammation with mild/severe interstitial edema, linear intra-alveolar fibrin deposition and early intra-alveolar organization ^6^. These findings provide an anatomopathological basis that could explain O_2_ diffusion failure and hypoxemia, revealing that acute inflammation is crucial for the severe progression of disease ^7^. This was further confirmed by the recent demonstration of reduced mortality in patients with severe COVID-19 who were treated with dexamethasone ^8^. Despite intensive efforts to find effective therapeutic strategies, mortality rates remain high, ranging from 5% to 49%, in patients admitted to hospitals and intense care units (ICUs) ^9-13^.

SARS-CoV-2, the pathogenic agent of COVID-19, enters human cells employing angiotensin converting enzyme 2 (ACE2) as a receptor ^14^. Lung alveolar epithelial cells and macrophages express high levels of ACE2 and thus are primary targets for SARS-CoV-2 ^15^. At least two studies have shown that, different from the influenza virus, the SARS-CoV-2 infection is accompanied by low expression levels of type I and II interferons and a massive production of inflammatory cytokines ^16,17^, suggesting this virus has an efficient immunomodulatory capacity that could explain, at least in part, its pathogenicity ^7^.

Physiologically, ACE2 catalyzes the reaction that inactivates the inflammatory substance DR9-bradykinin ^18^. In an experimental model of lung inflammation, the inhibition of ACE2 increased inflammatory cell recruitment and alveolar edema, whereas the inhibition of bradykinin protected the lungs from severe inflammatory damage ^18^. Following SARS-CoV-2 binding, ACE2 is internalized to endosomes, resulting in a subcellular shift that could affect its capacity to regulate bradykinin degradation ^17,19-22^. Here, we hypothesized that increased bradykinin levels could be one of the mediators of severe lung inflammation in COVID-19, and pharmacological inhibition of bradykinin could beneficially change the course of severe SARS-CoV-2 infection. A similar mechanism has been proposed for SARS-CoV lung damage ^23^.

In this proof-of-concept study, we evaluated 30 patients with severe COVID-19 that were randomized to one of the following treatments: standard care, icatibant (a bradykinin receptor 2 inhibitor) or inhibitor of C1 esterase/kallikrein (iC1e/K). The primary outcomes were death and time to clinical improvement. In this small cohort, we could not find evidence for a beneficial role of pharmacological inhibition of the kinin-kallikrein system in severe COVID-19 mortality and time to clinical improvement; however, both icatibant and iC1e/K promoted significant improvement of lung CT scores and increased blood eosinophil counts.

## Results

From April 23, 2020 to June 14, 2020, 241 patients were assessed for eligibility and 30 were included in the study (Fig. 1; Suppl. Tables 1 and 2). During the study period, the total number of patients screened accounted for 90% of all patients seeking medical attention at the Hospital & Clinics of the University of Campinas due to clinical suspicion of COVID-19. Patients included in the study represented 31% of all patients diagnosed with COVID-19 during this period. Supplementary Table 1 presents the reasons for the exclusion of screened patients. Supplementary Table 2 provides the chronological sequence of inclusion and randomization. Ten patients were randomized to each group; standard care, icatibant and iC1e/K.

**Figure 1.**
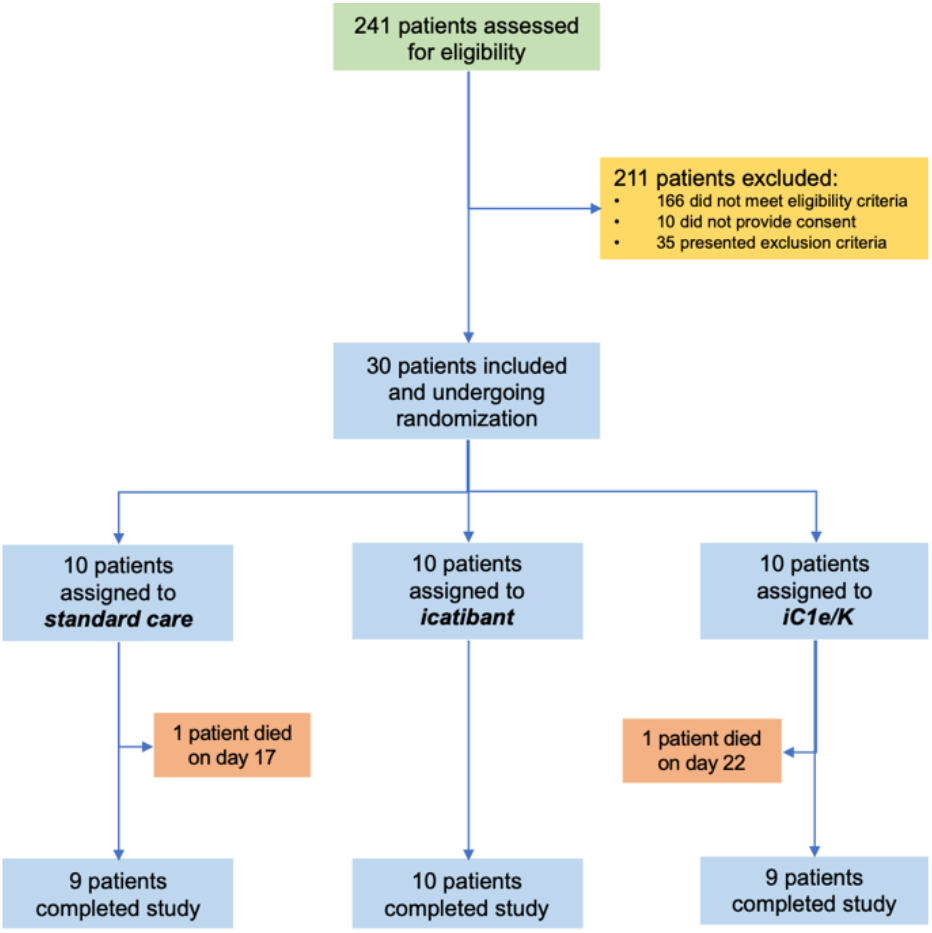
Schematic representation of patient screening, selection and randomization. iC1e/K, inhibitor of C1 esterase/kallikrein.

Table 1 presents the baseline parameters of all patients included in the study. The mean age was 51 years, 46% were women and the mean time to hospital admission from the onset of symptoms was eight days. The gender, age and demographics in each group were similar. The baseline body temperature, blood O_2_ saturation, lung CT score, systolic blood pressure, diastolic blood pressure, white blood cells, blood lymphocytes, blood platelets, plasma glucose, serum creatinine, aspartate aminotransferase (AST), lactate dehydrogenase (LDH), creatine kinase (CK), and C-reactive protein levels in each group were similar (Table 1). At baseline, alanine aminotransferase (ALT) was significantly higher in the icatibant group; nevertheless, all patients in this group had ALT levels within the inclusion range.

**Table 1.**
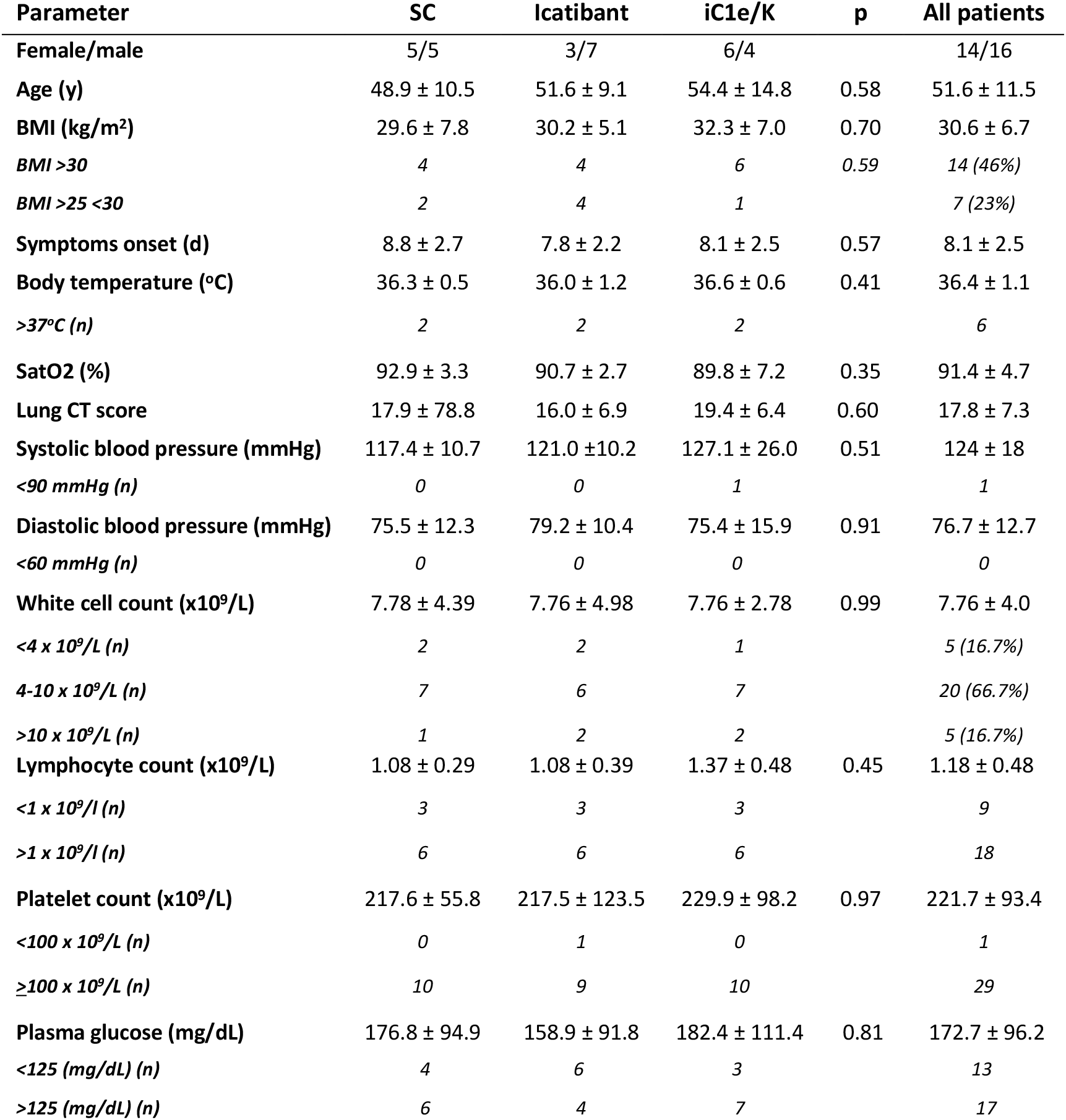
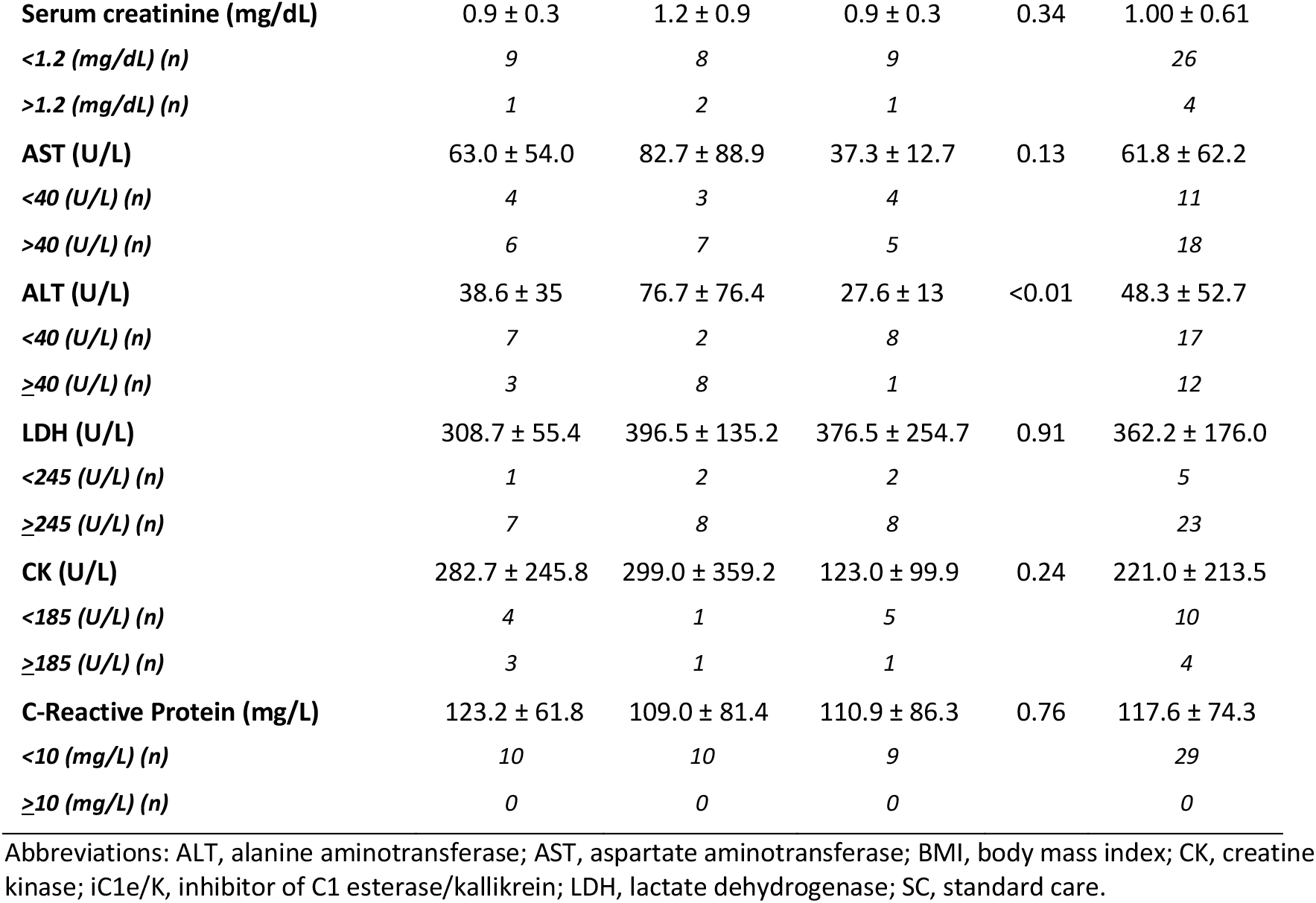
Base line parameters of patients included in the study.

Among the 30 patients included in the study, 25 were previously diagnosed with other medical conditions (Table 2). Hypertension was the leading comorbidity, affecting 50% of the patients. Diabetes and obesity were present in 47% and 43% of the patients, respectively. Overweight, dyslipidemia, hypothyroidism, asthma and fibromyalgia were also present in this cohort. Four patients were former smokers, whereas only one was currently smoking at the time of inclusion in the study (Table 2).

**Table 2.** Pre-morbid conditions of patients included in the study.

| Comorbidities at baseline | All patients<br>(n:30) | SC<br>(n:10) | Icatibant<br>(n:10) | iC1e/K<br>(n:10) | P-value |
| --- | --- | --- | --- | --- | --- |
| Hypertension, n (%) | 15 (50) | 4 (40) | 4 (40) | 7 (70) | 0.30 |
| Diabetes Mellitus, n (%) | 14 (46.7) | 5 (50) | 4 (40) | 5 (50) | 0.88 |
| Obesity, n (%) | 13 (43.3) | 3 (30) | 4 (40) | 6 (60) | 0.39 |
| Overweight, n (%) | 7 (23.3) | 2 (20) | 4 (40) | 1 (10) | -- |
| Dyslipidemia, n (%) | 5 (16.67) | 1 (10) | 1 (10) | 3 (30) | -- |
| Former smoker, n (%) | 4 (13.3) | 1 (10) | 1 (10) | 2 (20) | -- |
| Hypothyroidism, n (%) | 2 (6.67) | 0 (0) | 0 (0) | 2 (20) | -- |
| Asthma, n (%) | 1 (3.33) | 0 (0) | 0 (0) | 1 (10) | -- |
| Current smoker, n (%) | 1 | 1 (10) | 0 (0) | 0 (0) | -- |
| Fibromyalgia, n (%) | 1 (3.33) | 0 (0) | 1 (10) | 0 (0) | -- |

Abbreviations: iC1e/K, inhibitor of C1 esterase/kallikrein; SC, standard care.

Upon inclusion, all patients were diagnosed with severe COVID-19 based on SARS-CoV-2 diagnosis by RT-PCR, low blood O_2_ saturation levels (Table 1), high qSOFA (Fig. 2A) and high lung CT score (Table 1; Fig. 2B and Supplementary Figures 1-3); there were no significant differences in admission severity parameters among the groups.

**Figure 2.**
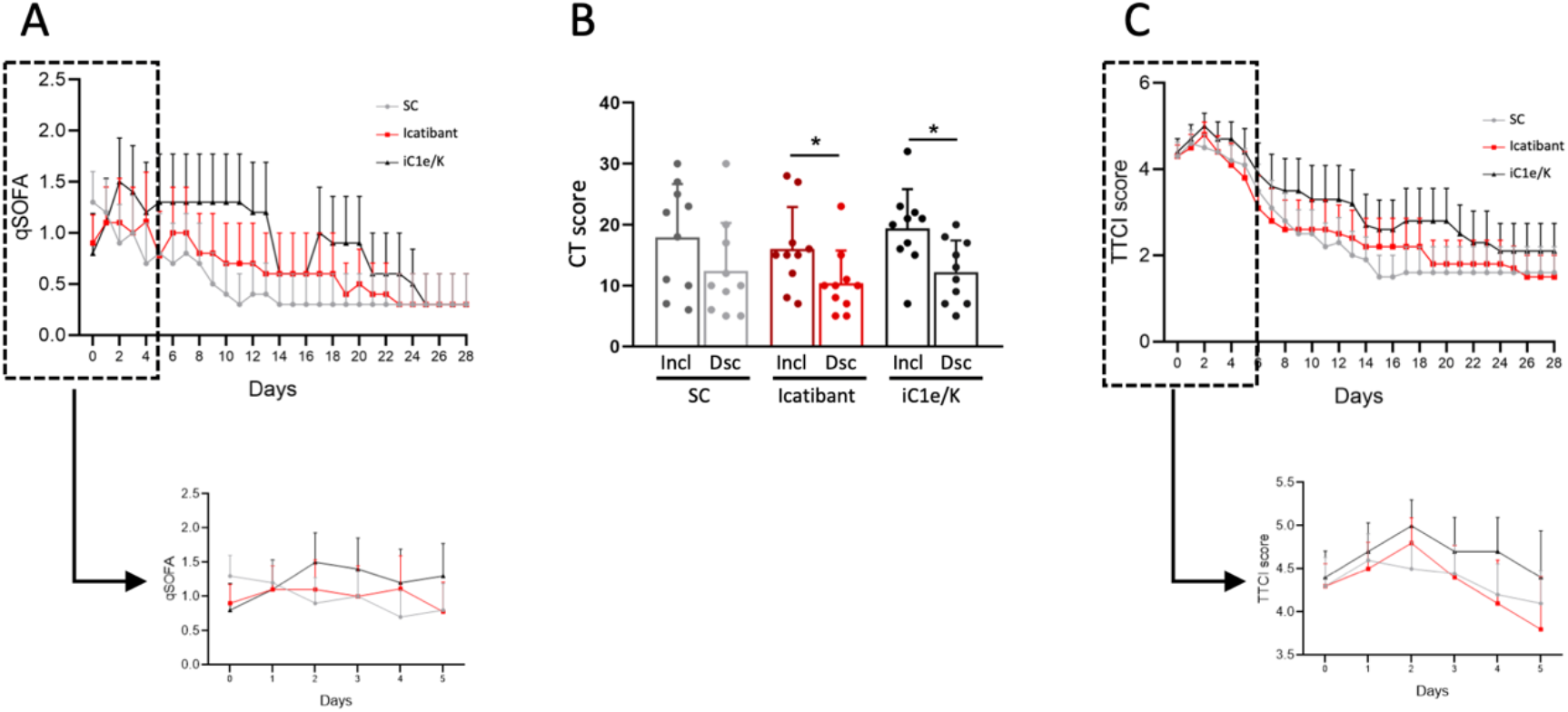
Major clinical and image-related outcomes. The quick sepsis-related organ failure score (qSOFA) was determined daily throughout hospitalization (A). Computed tomography lung score was obtained at inclusion and discharge (B). Time to clinical improvement (TTCI) was determined daily throughout hospitalization (C). Insets represent qSOFA (A) and TTCI (C) during the initial five days of treatment. *p<0.05. Dsc, discharge; iC1e/K, inhibitor of C1 esterase/kallikrein; Incl, inclusion; SC, standard care.

Neither icatibant nor iC1e/K pharmacological interventions significantly modified primary outcomes as compared to the standard care group (Table 3). Overall, there were two deaths, one in the standard care group (on day 17) and one in the iC1e/K group (on day 21). Times from admission to discharge were 10.5, 11.0 and 14.2 days in the standard care, icatibant and iC1e/K groups, respectively (p=0.62), whereas times in the ICU were 4.6, 6.2 and 8.7 days in the standard care, icatibant and iC1e/K groups, respectively (p=0.71). Clinical improvement was similar among the groups, as determined by the qSOFA (Fig. 2A) and TTCI score (Fig. 2C). This was also true when comparison was performed during the initial five days since inclusion in the study, which was the period when pharmacological intervention was undertaken (Fig. 2A, inset and Fig. 2C, inset). Nevertheless, both, icatibant and iC1e/K promoted significant reductions in lung CT scores (Fig. 2B).

**Table 3.** Primary outcomes of patients included in the study.

|  | SC | Icatibant | iC1e/K | p | All patients |
| --- | --- | --- | --- | --- | --- |
| TATD (days) | 10.5 ± 7.1 | 11.0 ± 8.9 | 14.2 ± 10.1 | 0.62 | 11.9 ± 8.6 |
| TIICU (days) | 4.6 ± 8.9 | 6.2 ± 10.5 | 8.7 ± 11.8 | 0.71 | 6.5 ± 10.2 |
| Deaths | 1 | 0 | 1 |  | 2 |

Abbreviations: iC1e/K, inhibitor of C1 esterase/kallikrein; TATD, time from admission to discharge; TIICU

Neither icatibant nor iC1e/K pharmacological interventions significantly modified blood immune cell counts as compared to the standard care group (Fig. 3A-3E and Suppl. Table 3). As compared to inclusion, the discharge white blood cell counts were unchanged in all three groups (Fig. 3A, Suppl. Table 3), lymphocytes were increased in the standard care and icatibant groups (Fig. 3B, Suppl. Table 3), neutrophils were unchanged in all three groups (Fig. 3C, Suppl. Table 3), monocytes were increased in the iC1e/K group (Fig. 3D, Suppl. Table 3) and eosinophils were increased in the icatibant and iC1e/K groups (Fig. 3E, Suppl. Table 3). The intervention did not modify the serum levels of IL-1β (Fig. 3F), whereas IL-6 was reduced in all three groups (Fig. 3G).

**Figure 3.**
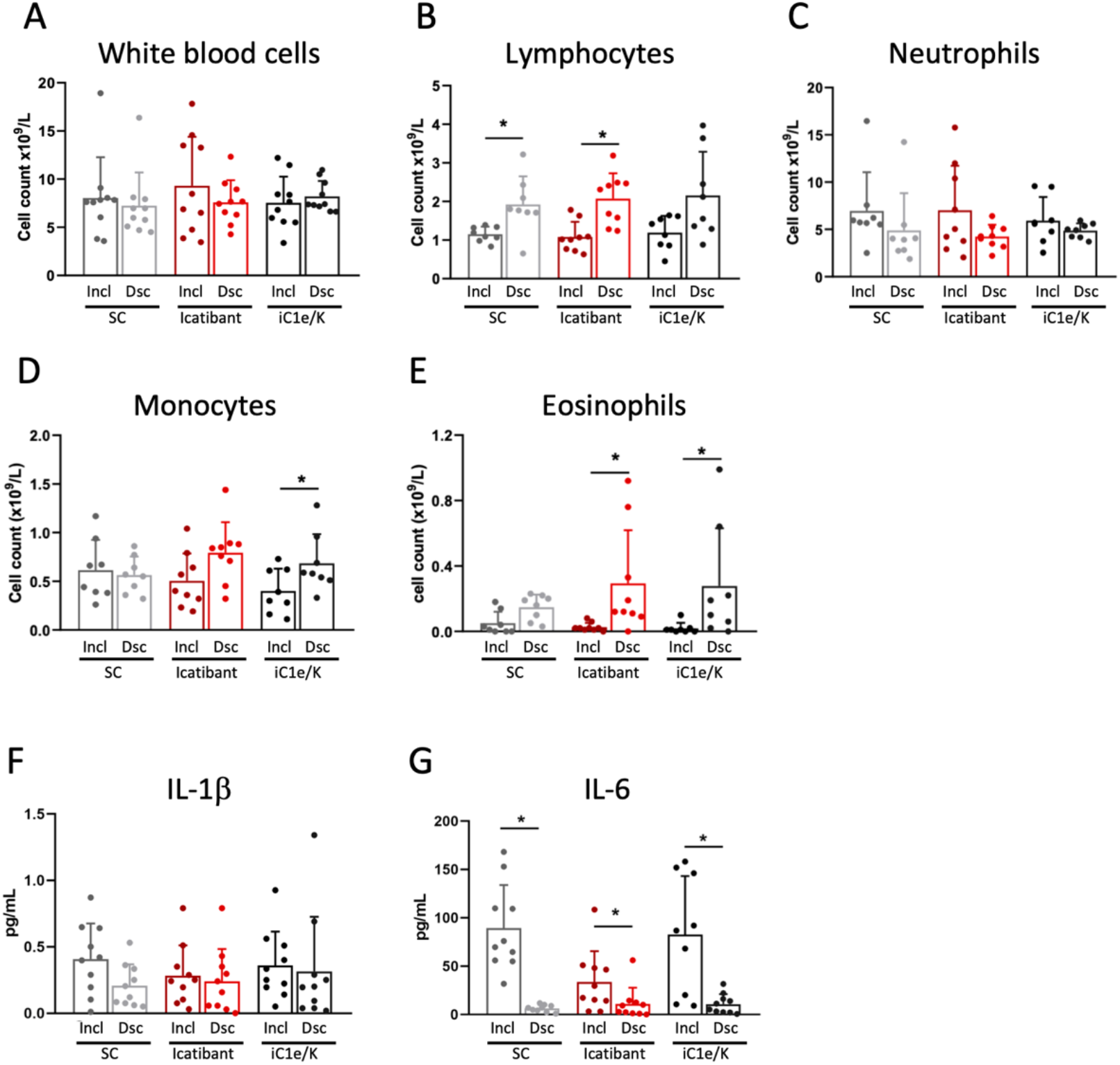
Blood immune cell and cytokine outcomes. White blood cells (A), lymphocytes (B), neutrophils (C), monocytes (D) and eosinophils (E) were determined by automated method in blood samples collected at inclusion and discharge. Interleukin-1 beta (F) and interleukin-6 were determined using ELISA kits in serum samples collected at inclusion and discharge. Dsc, discharge; iC1e/K, inhibitor of C1 esterase/kallikrein; IL-1b, interleukin-1 beta; IL-6, interleukin-6; Incl, inclusion; SC, standard care. *p<0.05.

At baseline, parameters related to the coagulation system were similar among all three groups (Suppl. Table 4). Neither icatibant nor iC1e/K pharmacological interventions significantly modified any of the coagulation parameters evaluated in this study (Fig. 4, Suppl. Table 4). As compared to inclusion, at discharge, blood platelets were increased in all three groups (Fig. 4A; Suppl. Table 4), whereas D-dimer (Suppl. Table 4), prothrombin time (Fig. 4B; Suppl. Table 4) and partial thromboplastin time (Fig. 4C; Suppl. Table 4) remained unchanged. There were no significant changes in blood levels of urea and creatinine (Suppl. Table 5).

During the intervention, there were no significant differences in reported adverse effects among the groups (Suppl. Table 6). As a whole, 14 patients presented with increased AST/ALT, five patients presented with diarrhea, three patients presented with nausea, three patients presented with vomiting, three patients presented with increased blood bilirubin, two patients presented with bradycardia and none of the patients presented with cardiac arrhythmias. No additional adverse effects were reported in the intervention groups.

**Figure 4.**
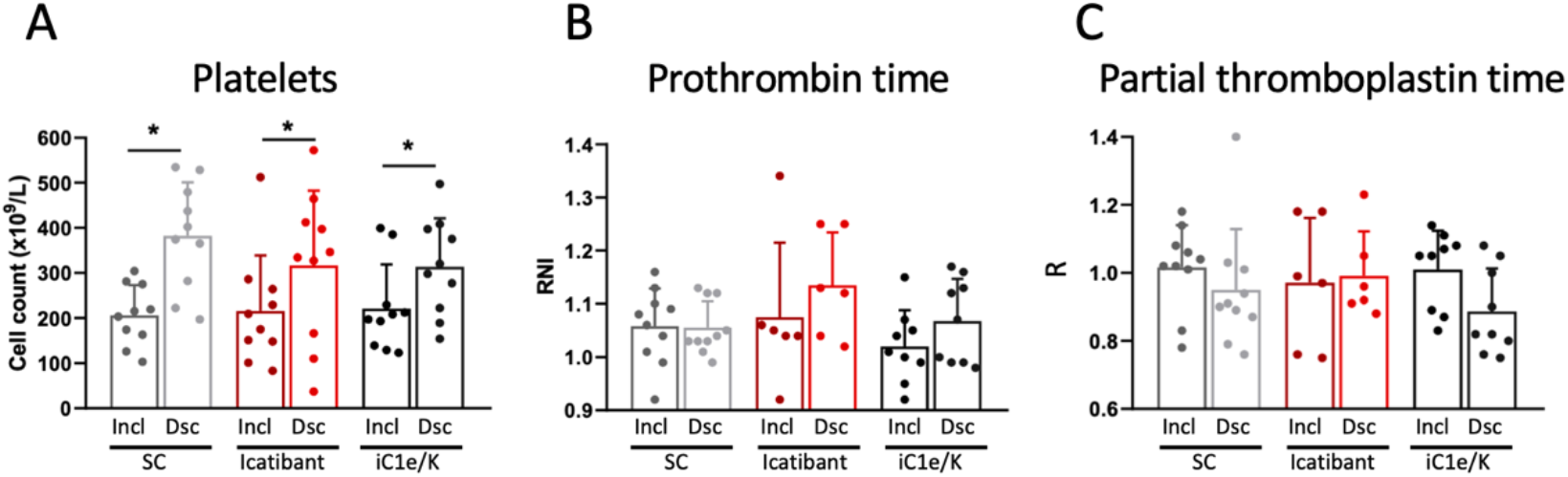
Coagulation-related parameters. Blood platelet counts (A), prothrombin time (B) and partial thromboplastin time (C) were determined in blood samples collected at inclusion and discharge. Dsc, discharge; iC1e/K, inhibitor of C1 esterase/kallikrein; Incl, inclusion; SC, standard care. *p<0.05.

## Discussion

In this proof-of-concept, randomized clinical trial, we could find neither a reduction of mortality nor a reduction of time to clinical improvement in a small cohort of patients with severe COVID-19 treated with pharmacological inhibitors of the kinin-kallikrein system. Nevertheless, there were significant reductions of lung CT scores, which could suggest a positive impact of pharmacological intervention on lung recovery, and increases in blood eosinophils, which is regarded as an indicator of disease recovery ^24^. In addition, interventions promoted no increases in the incidence of adverse effects, suggesting the drugs are safe for use in patients with severe COVID-19.

As for other emerging viral diseases, development of vaccine and effective antiviral agents are expected to provide maximum advances in prevention and therapeutics ^25^. However, even in the face of the current global efforts to develop such strategies, years may pass before successful therapies are available for worldwide use ^26,27^. Thus, drug repurposing has emerged as a potentially useful strategy to gain time and promote some advance in the battle against SARS-CoV-2^28^; in this scenario, existing antiviral drugs are undisputedly the natural candidates, and a number have been tested so far^29^. Unfortunately, clinical trials with lopinavir-ritonavir and remdesivir resulted in no change in mortality or severity of disease ^11,12^, whereas the combination of interferon beta-1b, lopinavir-ritonavir and ribavirin resulted only in shorter median time to negative nasopharyngeal swab, suggesting viral shedding could be abbreviated^30^.

As proposed elsewhere, drugs that act in distinct stages of the SARS-CoV-2 cell entrance and replication cycle could promote beneficial outcomes in the treatment of COVID-19 ^29^. Due to its action as a SARS-CoV-2 cell entry receptor, ACE2 has emerged as a primary candidate for pharmacological approaches aimed at reducing viral entrance and mitigating disease severity ^31,32^. It has been suggested that, in addition to mediating viral entrance, ACE2-SARS-CoV-2 physical interaction could modify ACE2 catalytic activity, impacting on the regulation of three independent (but rather integrated) systems—bradykinin, angiotensin and coagulation ^21,33-35^. This could explain, at least in part, the development of the most lethal triad in COVID-19 (i.e., severe lung inflammation, acute cardiovascular failure and thromboembolic events).

Here, we hypothesized that pharmacological inhibition of the kinin-kallikrein system could alleviate the acute inflammatory response during the initial steps of SARS-CoV-2 infection and thus provide a therapeutic advance against COVID-19. We used two drugs currently approved for the treatment of hereditary angioedema, icatibant and iC1e/K, which were compared against standard care. We chose standard care alone as the comparator because, when the trial began, there was no evidence for a pharmacological approach that could impact positively on COVID-19. Currently, there is at least one study showing a positive impact for dexamethasone reducing mortality in severe COVID-19 ^8^, and further studies with inhibitors of the kinin-kallikrein system should consider adding dexamethasone to all groups. Placebo and blinding were not adopted because of the urgent nature of this medical condition, and this was approved by the ethical committee.

Icatibant is an inhibitor of bradykinin receptor 2, which is approved for the treatment of bradykinin-induced edema in hereditary angioedema, leading to rapid relief of symptoms ^36^. Side effects are rare and usually restricted to inflammatory signals in the site of injection ^37,38^. In hereditary angioedema, a single 30 mg dose of icatibant is capable of reducing symptoms in minute to a few hours ^38^. As icatibant has a short half-life (6 h) ^39^, we used a 30 mg/dose every 8 h for four days, which proved safe, as no increased adverse effects were reported.

iC1e/K is a human-derived protein with an excellent record of beneficial actions in hereditary angioedema ^40,41^. It is well tolerated by most patients, and adverse effects are usually restricted to rash and headache ^42^. Due to its long half-life (56 h), we used 20 IU/kg at inclusion and on day four. There was no increased appearance of adverse effects in our cohort. It is worthwhile to mention that iC1e/K is present in human plasma, and ongoing clinical studies employing convalescent plasma to treat COVID-19 ^43^ may benefit not only from the presence of immunoneutralizing antibodies against SARS-CoV-2 but also from its endogenous capacity to inhibit the kinin-kallikrein system, as shown for hereditary angioedema ^44,45^.

The decision to pharmacologically intervene during the first four days after patient enrolment was based on the fact that bradykinin is a fast-acting, acute-phase inflammatory substance that has a very short half-life and is promptly produced upon exposure to the triggering agent ^46,47^. We reasoned that, if viral interaction could modify ACE2 catalytic activity, bradykinin would accumulate during the initial steps of COVID-19 progression and pharmacological inhibition would produce clinical outcomes only if used early after infection. Unfortunately, as in all other clinical reports on COVID-19 ^2,8,11,12^, in this study, the mean time since symptom appearance to hospital admittance was eight days, which could be somewhat late for intervention upon the bradykinin system. This could be one of the reasons explaining why we did not find a reduction in time to clinical improvement in our patients.

Even though interventions were incapable of modifying mortality and time to clinical improvement, the reduction of lung CT scores and increased blood eosinophils could indicate a beneficial impact of intervention on patient recovery. As recently reported, the mean time for poorest lung CT scores, which is defined as peak-stage or stage-3, is 9–13 days since beginning of symptoms ^48^. Thus, most patients included in our study were about to enter the time window when stage-3 develops. It is currently unknown if severe COVID-19 acute lung lesions could promote long-term structural abnormalities and, if so, how could it impact on respiratory function. In at least two studies, lung CT abnormalities were shown to persist after four weeks since the beginning of disease ^48,49^. Thus, it is intuitive to believe that improved lung CT scores in patients treated with kinin-kallikrein inhibitors indicate lung recovery that could impact on long-term respiratory function. If future studies show late respiratory loss in patients with COVID-19, it would be interesting to re-evaluate this cohort using functional respiratory tests.

The increased blood eosinophils provide yet another indication of improved recovery in the groups undergoing pharmacological intervention. In fatal cases of COVID-19, blood eosinophil counts were consistently reduced at hospital admittance ^50^, whereas autopsies revealed an absence of eosinophils in lung specimens ^51^. In addition, a low blood eosinophil count was identified as a risk factor (odds ratio = 10.2) for severe progression of COVID-19 ^52^, whereas the progressive increase of blood eosinophils was associated with disease recovery ^24^. Recent studies have challenged the classic concept of eosinophil roles being restricted to helminthic parasitosis and allergic diseases ^53^. Eosinophils have been shown to act centrally in immunomodulatory networks that warrant homeostasis during inflammatory responses ^54^ and also to promote antiviral immunity in influenza A virus infection ^55^. This has been proposed as the mechanism underlying the protective impact of asthma on influenza morbidity ^55^, which could also be applied to COVID-19 (doi: 10.22541/au.159118771.11841404).

The main weaknesses of this study are the small number of patients, and the fact that pharmacological intervention was instituted approximately eight days after the beginning of symptoms. An important reason for restricting the cohort to 30 patients was that, during enrollment, it was demonstrated that dexamethasone could reduce mortality in patients with severe COVID-19 ^8^. Thus, we consider that the continuity of ongoing protocols or beginning of new protocols should ethically include dexamethasone in all arms. Conversely, regarding the fact that the mean time for patient inclusion was eight days, which could be a late stage for interventions targeting bradykinin, there is little that could be done, because of the natural course of the disease, which prompts patients to seek medical attention at this stage. Even if patients were advised to seek medical attention at an earlier stage, most of them would not progress to severe COVID-19. Therefore, only the development of methods that predict severe progression of disease at an early stage could reduce time to inclusion.

In conclusion, this is the first clinical trial reporting the impact of pharmacological inhibition of the kinin-kallikrein system in patients with severe COVID-19. The lack of findings regarding mortality and time to clinical improvement could be attributed to small sample size and beginning the intervention at a late stage. Nevertheless, improvement in CT lung scores and increased blood eosinophils suggest that intervention had a beneficial impact on patient recovery and should be considered in future trials.

## Methods

### Study design and patients

This was a phase II, single center, three-arm parallel group, open-label randomized clinical trial (RCT) conducted at the Clinics Hospital, University of Campinas. Enrollment occurred from April 22, 2020 to June 14, 2020. There was a 1:1:1 allocation ratio to icatibant plus standard care *vs*. iC1e/K plus standard care *vs*. standard care alone. The study followed the principles of the Declaration of Helsinki. Inclusion criteria were age at least 18 years at the time of signing the consent form, symptom duration of 12 days or less upon recruitment, SARS-CoV-2 diagnosis by RT-PCR method according to the Berlin-Charité protocol ^56^, diagnosis of COVID-19 typical pneumonia confirmed by CT of the chest and scored by two lung expert radiologists, SpO_2_ ≤ 94% in ambient air or Pa0_2_/FiO_2_ ≤ 300 mmHg, willingness of study participants to accept randomization to any assigned treatment arm, patient or responsible family member signing the consent form, and agreement that patient should not enroll in any other experimental study prior to completion of the 28-day follow-up. Exclusion criteria were pregnant or breastfeeding women (a beta-HCG level was determined in all eligible women of childbearing age); severe renal impairment (estimated glomerular filtration rate ≤ 30 ml/min/1.73 m^2^), patients receiving continuous renal replacement therapy (hemodialysis or peritoneal dialysis), or previous renal transplant; severe liver disease (AST or ALT 5X above the reference value); HIV infection (HIV was tested for in all eligible patients); or patients with any other immunodeficiencies, previous diagnosis of cancer, previous diagnosis of hereditary angioedema, previous ischemic myocardial disease, previous thromboembolic disease, current use of immunosuppressive drug therapy, or use of any experimental treatment for SARS-CoV-2 infection within 30 days prior to screening. This trial has been registered at the Brazilian Clinical Trials Registry (http://www.ensaiosclinicos.gov.br/rg/?q=U1111-1250-1843); Universal Trial Number (UTN): U1111-1250-1843 and approved by the local Ethical Review Committee of the Clinics Hospital of the University of Campinas (protocol CAEE: 30227920.9.0000.5404). Written consent to participate in this study has been obtained from all patients or substitute decisionmakers (for patients lacking decision-making capacity).

### Randomization

The randomization list was generated by an independent researcher with no involvement in the study. A software-based approach (www.sealedenvelope.com) was used to generate the allocation sequence. Sealed and sequentially numbered opaque envelopes, containing the allocation arms, were prepared in advance according to the randomization list. The project coordinator and investigators in each unit of the hospital enrolled the participants. Once the eligibility criteria were met, the envelope was opened, and the therapy was prescribed according to the arm specified in the envelope.

### Blinding

This was an open-label trial; thus, neither the trial participant nor the care providers were blinded. However, data entry staff and data manager were blinded. Data was sent to the data entry staff by code, and the study arm was never specified. Data entry staff and data managers were not part of the hospital staff and were never given access to any clinical data belonging to the patients. The Data Safety Monitoring Board (DSMC) was unblinded.

### Procedures

Group A, Firazyr® (icatibant acetate 30 mg [3.0 ml of 10 mg/ml solution]) subcutaneous injections in the abdominal area were administered at intervals of 8 h for 4 days *plus* standard care. Group B, Berinert® (human plasma-derived C1 esterase/kallikrein Inhibitor) administered at a dose of 20 IU/kg body weight on day 1 shortly after recruitment and on day 4 (each vial contains 500 IU of C1 esterase/kallikrein inhibitor as a lyophilized product for reconstitution with 10 ml of sterile water for injection) *plus* standard care. Group C, standard care. Upon medical decision, patients received antibiotics, antithrombotic therapy, oxygen support, non-invasive and invasive mechanical ventilation, vasopressor drugs, stress doses of corticosteroids and renal support therapy. The criteria for eventual discontinuation of the intervention were withdrawal of consent, grade 4 adverse reaction or allergy to the drug, and protocol violation.

### Clinical, radiological and laboratory monitoring

Diagnosis of SARS-CoV-2 infection was performed by RT-PCR in nasopharyngeal swab samples. Once patients were included in the study, clinical data was obtained continuously and uploaded once a day on the data management system. CT scans were performed at admittance and at days 14 and 28, or otherwise at hospital discharge. CT scan scoring was performed by two experienced radiologists and followed guidelines published elsewhere ^48^. Blood samples were collected on admittance and at days 7, 14, 21 and 28, or otherwise at hospital discharge. Additional clinical, radiological, and laboratory tests were performed at any time at the discretion of the medical staff. Hematological and biochemical parameters were determined in blood samples using automated methods. Cytokines were determined using ELISA kits, according to the recommendations provided by the manufacturers (Interleukin-1beta, RD Systems – catalog number HSLB00C; interleukin-6, RD Systems – catalog number D6050).

### Data management

Each patient allocated in the study was given a unique code generated by the independent researcher who created the randomization list. The codes were stored in a password protected file that was not accessible to the data management team or other investigators. All data (outcomes, questionnaires, clinical data, CT, and laboratory measurements) were retrieved from the online official medical records of the hospital and were immediately uploaded into REDCap (https://redcap.vanderbilt.edu). Data were retrieved daily by one investigator using a datasheet available online and in print form. Collected data were sent by email to two researchers on the data management team in order to establish double data entry.

### Outcomes

The primary outcome was the time to clinical improvement (TTCI) as defined by the Cap-China Network ^57^, which was employed in the LOTUS China trial ^11^ and recommended by the World Health Organization. TTCI refers to the time from randomization to an improvement of two points on the seven-category ordinal scale as follows ^58^ (or discharge alive from the hospital): 1, not hospitalized with resumption of normal activities; 2, not hospitalized, but unable to resume normal activities; 3, hospitalized, not requiring supplemental oxygen; 4, hospitalized, requiring supplemental oxygen; 5, hospitalized, requiring nasal high-flow oxygen therapy, non-invasive mechanical ventilation; 6, hospitalized, requiring ECMO, invasive mechanical ventilation, or both; or 7, death. The secondary outcomes were as follows: percentage of patients on seven-category scale on days 7, 14 and 21; time on mechanical ventilation; time of hospitalization of survivors; time on oxygen support; time from randomization to hospital discharge; time from randomization to death; and frequency of severe adverse events as defined by National Cancer Institute Common Terminology Criteria for Adverse Events (CTCAE), version 5.0. Quick sepsis-related organ failure score (qSOFA) was determined as defined elsewhere (qsofa.org).

### Statistical analysis

All statistical analyses were performed using SPSS version 22.0 and GraphPad Prism 8.0. Values are presented as means and standard deviation in tables and as means and standard error of the mean in figures. To check the normal distribution of variables, the Kolmogorov-Smirnov test was used. When the variable did not present a normal distribution, the equivalent non-parametric test was used. For comparisons between the three groups at baseline, the unidirectional analysis of variance (ANOVA) was used. For non-parametric variables, the Kruskal-Wallis test was employed. Also, a paired Student’s *t* test (or Wilcoxon test) was used to compare variables at admission and at discharge for all three groups. To test for differences in frequencies between groups, we used the chi-square test. A p-value of less than 0.05 was considered statistically significant.

## Data Availability

Data are available upon request

## Author contribution

LAV is the supervisor and lead contact. LAV conceived the study, raised funds, organized the protocol and wrote the paper. EM coordinated the execution team and provided thoughtful insights for the design of the study. MLM was the cosupervisor of the study. ACP, RGU, LCR, AFB, TAN and MVA screened, enrolled and followed-up patients. BB, RLC and ASF performed laboratory analysis. MM-P and EPA organized data. ALEF, TMS, PT and MLM provided clinical supervision support for the preparation and execution of the trial. RPS and SSD designed and performed radiological analysis. MM-P performed statistical analysis. All authors read and approved the final manuscript.

## Data sharing

All data obtained in this study can be made available to qualified researchers upon request, provided that data protection and ethical standards are in compliance with the principles of the Declaration of Helsinki.

## Grant providers

Sao Paulo Research Foundation grants #2013/07607-8 and #2020/04522-5; University of Campinas grant #2298/20.

---

**Supplementary Table 1.** Patients screened and excluded from study.

| Excluded Patients | Age | Gender | Exclusion Criteria |
| --- | --- | --- | --- |
| E1 | 50-60 | M | Symptoms >12 days |
| E2 | 60-70 | F | Symptoms >12 days |
| E3 | 50-60 | M | Symptoms >12 days |
| E4 | 40-50 | M | Symptoms >12 days |
| E5 | 70-80 | F | Symptoms >12 days |
| E6 | 50-60 | M | Symptoms >12 days |
| E7 | 70-80 | F | Symptoms >12 days |
| E8 | 40-50 | M | Symptoms >12 days |
| E9 | 60-70 | F | Symptoms >12 days |
| E10 | 50-60 | M | Symptoms >12 days |
| E11 | 60-70 | M | Symptoms >12 days |
| E12 | 70-80 | F | Symptoms >12 days |
| E13 | 50-60 | F | Symptoms >12 days |
| E14 | 40-50 | M | Sat >94% |
| E15 | 40-50 | M | Sat >94% |
| E16 | 40-50 | M | Sat >94% |
| E17 | 20-30 | M | Sat >94% |
| E18 | 50-60 | M | Sat >94% |
| E19 | 30-40 | F | Sat >94% |
| E20 | 60-70 | F | Sat >94% |
| E21 | 30-40 | M | Sat >94% |
| E22 | 30-40 | M | Kidney transplant |
| E23 | 50-60 | M | Kidney transplant |
| E24 | 50-60 | M | Kidney transplant |
| E25 | 30-40 | F | Kidney transplant |
| E26 | 18-20 | F | Kidney transplant |
| E27 | 40-50 | F | Kidney transplant |
| E28 | 90-100 | M | DNPC |
| E29 | 80-90 | M | DNPC |
| E30 | 90-100 | M | DNPC |
| E31 | 80-90 | M | DNPC |
| E32 | 80-90 | F | DNPC |
| E33 | 50-60 | M | DNPC |
| E34 | 20-30 | M | DNPC |
| E35 | 60-70 | M | DNPC |
| E36 | 50-60 | M | DNPC |
| E37 | 60-70 | M | DNPC |
| E38 | 60-70 | M | CKD |
| E39 | 50-60 | F | CKD |
| E40 | 60-70 | F | CKD |
| E41 | 50-60 | M | CKD |
| E42 | 50-60 | F | HIV |
| E43 | 20-30 | M | HIV |
| E44 | 50-60 | F | HIV |
| E45 | 60-70 | M | AMI |
| E46 | 70-80 | M | AMI |
| E47 | 50-60 | F | AMI |
| E48 | 50-60 | F | AMI |
| E49 | 50-60 | M | Hydroxychloroquine use |
| E50 | 50-60 | F | Hydroxychloroquine use |
| E51 | 40-50 | M | Hydroxychloroquine use |
| E52 | 20-30 | M | Systemic sclerosis |
| E53 | 60-70 | F | Systemic sclerosis |
| E54 | 60-70 | M | Stroke |
| E55 | 80-90 | M | Stroke |
| E56 | 40-50 | M | Thromboembolic event |
| E57 | 60-70 | M | Pulmonary thromboembolism |
| E58 | 50-60 | M | Hepatic cirrhosis |
| E59 | 40-50 | M | Liver transplant |
| E60 | 40-50 | F | Autoimmune hepatitis |
| E61 | 50-60 | M | AML |
| E62 | 40-50 | M | ALL |
| E63 | 60-70 | M | Myasthenia gravis |
| E64 | 50-60 | M | FSGS |
| E65 | 20-30 | M | Polytrauma |
| E66 | 60-70 | F | Expansive brain injury |
| E67 | 70-80 | F | ND |
| E68 | 30-40 | F | ND |
| E69 | 60-70 | F | ND |
| E70 | 20-30 | M | ND |
| E71 | 70-80 | F | ND |
| E72 | 40-50 | M | ND |
| E73 | 50-60 | F | ND |
| E74 | 80-90 | M | ND |
| E75 | 60-70 | F | ND |
| E76 | 50-60 | M | ND |
| E77 | 50-60 | F | ND |
| E78 | 50-60 | F | ND |
| E79 | 60-70 | F | ND |
| E80 | 30-40 | M | ND |
| E81 | 60-70 | F | ND |
| E82 | 60-70 | F | ND |
| E83 | 50-60 | F | ND |
| E84 | 30-40 | F | ND |
| E85 | 18-20 | F | ND |
| E86 | 70-80 | F | ND |
| E87 | 30-40 | M | ND |
| E88 | 50-60 | F | ND |
| E89 | 20-30 | M | ND |
| E90 | 70-80 | M | ND |
| E91 | 60-70 | M | ND |
| E92 | 90-100 | F | ND |
| E93 | 50-60 | F | ND |
| E94 | 50-60 | F | ND |
| E95 | 40-50 | F | ND |
| E96 | 20-30 | M | ND |
| E97 | 50-60 | M | ND |
| E98 | 60-70 | M | ND |
| E99 | 40-50 | M | ND |
| E100 | 80-90 | M | ND |
| E101 | 20-30 | M | ND |
| E102 | 50-60 | M | ND |
| E103 | 50-60 | M | ND |
| E104 | 90-100 | M | ND |
| E105 | 70-80 | F | ND |
| E106 | 20-30 | F | ND |
| E107 | 60-70 | M | ND |
| E108 | 50-60 | M | ND |
| E109 | 60-70 | F | ND |
| E110 | 30-40 | M | ND |
| E111 | 70-80 | F | ND |
| E112 | 70-80 | M | ND |
| E113 | 80-90 | M | ND |
| E114 | 70-80 | M | ND |
| E115 | 60-70 | M | ND |
| E116 | 60-70 | M | ND |
| E117 | 30-40 | F | ND |
| E118 | 80-90 | F | ND |
| E119 | 50-60 | M | ND |
| E120 | 30-40 | F | ND |
| E121 | 40-50 | F | ND |
| E122 | 60-70 | M | ND |
| E123 | 30-40 | M | ND |
| E124 | 70-80 | M | ND |
| E125 | 60-70 | F | ND |
| E126 | 60-70 | M | ND |
| E127 | 60-70 | F | ND |
| E128 | 50-60 | M | ND |
| E129 | 20-30 | F | ND |
| E130 | 70-80 | M | ND |
| E131 | 70-80 | F | ND |
| E132 | 20-30 | M | ND |
| E133 | 30-40 | M | ND |
| E134 | 50-60 | F | ND |
| E135 | 70-80 | F | ND |
| E136 | 30-40 | M | ND |
| E137 | 60-70 | M | ND |
| E138 | 20-30 | M | ND |
| E139 | 70-80 | M | ND |
| E140 | 40-50 | M | ND |
| E141 | 40-50 | M | ND |
| E142 | 50-60 | M | ND |
| E143 | 80-90 | M | ND |
| E144 | 60-70 | M | ND |
| E145 | 80-90 | M | ND |
| E146 | 30-40 | F | ND |
| E147 | 60-70 | M | ND |
| E148 | 70-80 | M | ND |
| E149 | 60-70 | M | ND |
| E150 | 50-60 | F | ND |
| E151 | 40-50 | F | ND |
| E152 | 40-50 | M | ND |
| E153 | 60-70 | M | ND |
| E154 | 50-60 | M | ND |
| E155 | 70-80 | M | ND |
| E156 | 70-80 | F | ND |
| E157 | 60-70 | M | ND |
| E158 | 50-60 | M | ND |
| E159 | 50-60 | F | ND |
| E160 | 50-60 | M | ND |
| E161 | 60-70 | M | ND |
| E162 | 50-60 | M | ND |
| E163 | 50-60 | M | ND |
| E164 | 60-70 | M | ND |
| E165 | 60-70 | M | ND |
| E166 | 60-70 | M | ND |
| E167 | 20-30 | M | ND |
| E168 | 30-40 | F | ND |
| E169 | 80-90 | F | ND |
| E170 | 60-70 | M | ND |
| E171 | 70-80 | M | ND |
| E172 | 60-70 | M | ND |
| E173 | 60-70 | M | ND |
| E174 | 70-80 | F | ND |
| E175 | 50-60 | M | ND |
| E176 | 80-90 | F | ND |
| E177 | 70-80 | F | ND |
| E178 | 80-90 | M | ND |
| E179 | 50-60 | M | ND |
| E180 | 40-50 | F | ND |
| E181 | 50-60 | F | ND |
| E182 | 50-60 | M | ND |
| E183 | 40-50 | F | ND |
| E184 | 40-50 | M | ND |
| E185 | 50-60 | M | ND |
| E186 | 50-60 | M | ND |
| E187 | 80-90 | F | ND |
| E188 | 80-90 | F | ND |
| E189 | 60-70 | M | ND |
| E190 | 70-80 | M | ND |
| E191 | 70-80 | M | ND |
| E192 | 30-40 | F | ND |
| **E193** | 40-50 | M | ND |
| **E194** | 50-60 | F | ND |
| **E195** | 50-60 | M | ND |
| **E196** | 60-70 | M | ND |
| **E197** | 70-80 | M | ND |
| **E198** | 50-60 | F | ND |
| **E199** | 50-60 | F | ND |
| **E200** | 70-80 | F | ND |
| **E201** | 70-80 | F | ND |
| **E202** | 50-60 | F | ND |
| **E203** | 70-80 | M | ND |
| **E204** | 30-40 | M | ND |
| **E205** | 50-60 | M | ND |
| **E206** | 70-80 | F | ND |
| **E207** | 40-50 | M | ND |
| **E208** | 70-80 | F | ND |
| **E209** | 20-30 | M | ND |
| **E210** | 60-70 | F | ND |
| **E211** | 30-40 | F | ND |

Abbreviations: ALL, acute lymphoid leukemia; AMI, acute myocardial infarction; AML, acute myeloid leukemia; CKD, chronic kidney disease; DNPC, did not provide consent; FSGS, focal segmental glomerulosclerosis; HIV, diagnosis of HIV infection; ND, no confirmation of diagnosis by PCR.

**Supplementary Table 2.** Patients included in the study.

| **Patient Identity** | **Age** | **Gender** | **Inclusion week** | **Time since beginning of symptoms (days)** | **Randomization** | **Arm** |
| --- | --- | --- | --- | --- | --- | --- |
| **P1** | 50-60 | M | 17 | 7 | 001 YD6 | SC |
| **P2** | 50-60 | M | 17 | 8 | 002 ZE3 | Icatibant |
| **P3** | 70-80 | F | 18 | 4 | 003 510 | iC1e/K |
| **P4** | 30-40 | M | 18 | 10 | 004 SU3 | SC |
| **P5** | 40-50 | M | 18 | 8 | 005 DH1 | Icatibant |
| **P6** | 50-60 | M | 19 | 9 | 006 GK4 | Icatibant |
| **P7** | 40-50 | M | 19 | 10 | 007 OH1 | SC |
| **P8** | 40-50 | M | 20 | 6 | 008 IE5 | iC1e/K |
| **P9** | 30-40 | M | 20 | 4 | 009 ZL0 | Icatibant |
| **P10** | 50-60 | M | 21 | 11 | 010 UW5 | Icatibant |
| **P11** | 60-70 | F | 21 | 7 | 011 YB8 | iC1e/K |
| **P12** | 50-60 | M | 21 | 10 | 012 UU6 | iC1e/K |
| **P13** | 60-70 | F | 21 | 7 | 013 DU2 | SC |
| **P14** | 50-60 | M | 22 | 12 | 014 KL6 | SC |
| **P15** | 40-50 | F | 22 | 10 | 015 GS8 | SC |
| **P16** | 40-50 | M | 22 | 10 | 016 TL8 | Icatibant |
| **P17** | 50-60 | F | 22 | 7 | 017 TT4 | SC |
| **P18** | 40-50 | F | 22 | 11 | 018 DC5 | iC1e/K |
| **P19** | 30-40 | F | 22 | 11 | 019 MI3 | SC |
| **P20** | 60-70 | M | 22 | 8 | 020 VZ9 | iC1e/K |
| **P21** | 40-50 | M | 22 | 5 | 021 QR4 | Icatibant |
| **P22** | 30-40 | F | 23 | 3 | 022 FM6 | iC1e/K |
| **P23** | 40-50 | F | 23 | 7 | 023 WG1 | SC |
| **P24** | 30-40 | M | 23 | 10 | 024 CC2 | iC1e/K |
| **P25** | 40-50 | F | 24 | 8 | 025 VK0 | iC1e/K |
| **P26** | 60-70 | M | 24 | 5 | 026 RL1 | SC |
| **P27** | 70-80 | F | 24 | 10 | 027 ZR9 | iC1e/K |
| **P28** | 60-70 | F | 24 | 9 | 028 LQ8 | Icatibant |
| **P29** | 50-60 | F | 24 | 8 | 029 GG3 | Icatibant |
| **P30** | 50-60 | F | 24 | 6 | 030 QT1 | Icatibant |

Abbreviations: iC1e/K, inhibitor of C1 esterase/kallikrein; SC, standard care.

**Supplementary Table 3.** Hematimetric values.

|  | Standard Care |  |  | Icatibant |  |  | iC1e/K |  |  | Total sample |  |  |
| --- | --- | --- | --- | --- | --- | --- | --- | --- | --- | --- | --- | --- |
|  | Admission | Discharge | p | Admission | Discharge | p | Admission | Discharge | p | Admission | Discharge | p |
| **White blood cells (x10\*\*3/μL)#** | 7.78 ± 4.38 | 7.27 ± 3.49 | 0.96 | 7.76 ± 4.98 | 7.87 ± 2.37 | 0.88 | 7.76 ± 2.76 | 7.79 ± 1.65 | 0.96 | 7.76 ± 4.00 | 7.64 ± 2.53 | 0.88 |
| **Segmented neutrophils (x10\*\*3/μL)#** | 5.81 ± 4.74 | 4.74 ± 4.00 | 0.16 | 6.60 ± 3.61 | 4.24 ± 1.24 | 0.05 | 5.80 ± 2.18 | 5.0 ± 0.82 | 0.85 | 6.08 ± 3.03 | 4.66 ± 2.30 | 0.03 |
| **Lymphocytes (x10\*\*3/μL)#** | 1.15 ± 0.20 | 1.92 ± 0.73 | 0.02 | 1.08 ± 0.39 | 2.07±0.66 | 0.01 | 1.19 ± 0.43 | 2.16 ± 1.13 | 0.05 | 1.14 ± 0.35 | 2.05 ± 0.83 | 0.01 |
| **Monocytes (x10\*\*3/μL)#** | 0.61 ± 0.31 | 0.57± 0.19 | 0.62 | 0.50 ± 0.28 | 0.79 ± 0.31 | 0.07 | 0.40 ± 0.23 | 0.69± 0.30 | 0.03 | 0.51 ± 0.28 | 0.69 ± 0.28 | 0.03 |
| **Eosinophils (x10\*\*3/μL)#** | 0.05 ± 0.07 | 0.15 ± 0.08 | 0.08 | 0.02 ± 0.07 | 0.15 ± 0.08 | 0.01 | 0.02 ± 0.03 | 0.28 ± 0.35 | 0.02 | 0.03 ± 0.05 | 0.24 ± 0.28 | 0.01 |

Abbreviation: iC1e/K, inhibitor of C1 esterase/kallikrein; # Non-parametric test

**Supplementary Table 4.** Blood coagulation parameter values.

|  | Standard care |  |  | Icatibant |  |  | iC1e/K |  |  | Total sample |  |  |
| --- | --- | --- | --- | --- | --- | --- | --- | --- | --- | --- | --- | --- |
|  | Admission | Discharge | p | Admission | Discharge | p | Admission | Discharge | p | Admission | Discharge | p |
| **D-dimer** | 1070.8 ± 745.5 | 2345.0 ± 3928.9 | 0.26 | 834.4 ± 398.2 | 2429.6 ± 3181.0 | 0.14 | 8984.5 ± 24961.8 | 2753.4 ± 4085.7 | 0.72 | 3821.1 ± 14,942 | 2518.4 ± 3629.0 | 0.17 |
| **Platelets (x10\*\*3/μL)** | 206.3 ± 66.8 | 382.0 ± 119.0 | <0.01 | 215.8 ± 123.2 | 316.5 ± 165.8 | 0.02 | 221.4 ± 97.21 | 313.7 ± 107.64 | <0.01 | 214.5 ± 95.2 | 337.4 ± 132 | <0.01 |
| **PT (sec)** | 12.51 ± 0.86 | 12.36 ± 0.49 | 0.57 | 12.81 ± 1.73 | 13.40 ± 1.38 | 0.75 | 12.06 ± 0.83 | 12.6 ± 0.97 | 0.31 | 12.42 ± 1.11 | 12.69 ± 0.99 | 0.55 |
| **PT (%)** | 90.75 ± 12.88 | 90.87 ± 7.85 | 0.88 | 90.18 ± 18.26 | 80.25 ± 12.38 | 0.46 | 97.49 ± 12.57 | 89.37 ± 12.52 | 0.21 | 93.04 ± 14.00 | 87.78 ± 11.24 | 0.28 |
| **PT (RNI)** | 1.06 ± 0.07 | 1.06 ± 0.05 | 0.92 | 1.07 ± 0.14 | 1.14 ± 0.10 | 0.50 | 1.02 ± 0.07 | 1.06 ± 0.08 | 0.21 | 1.05 ± 0.09 | 1.08 ± 0.08 | 0.27 |
| **PTT (sec)** | 29.23 ± 3.57 | 27.39 ± 5.15 | 0.31 | 27.61 ± 5.08 | 28.03 ± 3.70 | 0.74 | 29.09 ± 3.28 | 25.56 ± 3.65 | 0.07 | 28.75 ± 3.83 | 26.93 ± 4.27 | 0.07 |
| **PTT - R** | 1.02 ± 0.12 | 0.95 ± 0.18 | 0.31 | 0.97 ± 0.19 | 0.99 ± 0.13 | 0.91 | 1.01 ± 0.11 | 0.89 ± 0.13 | 0.07 | 1.00 ± 0.13 | 0.94 ± 0.15 | 0.07 |

Abbreviations: iC1e/K, inhibitor of C1 esterase/kallikrein; PT, sss; PTT, .

**Supplementary Table 5.** Parameters related to renal function.

|  | Standard care |  |  | Icatibant |  |  | iC1e/K |  |  | Total sample |  |  |
| --- | --- | --- | --- | --- | --- | --- | --- | --- | --- | --- | --- | --- |
|  | Admission | Discharge | p | Admission | Discharge | p | Admission | Discharge | p | Admission | Discharge | p |
| **Creatinine (mg/dL)** | 0.9 ± 0.3 | 1.0 ± 1.0 | 0.44 | 1.2 ± 1.0 | 1.6 ± 2.3 | 0.51 | 0.9 ± 0.3 | 0.8 ± 0.4 | 0.39 | 1.0 ± 0.6 | 1.1 ± 1.5 |  |
| **Urea (mg/dL)** | 37.6 ± 20.0 | 38.4 ± 33.5 | 0.94 | 40.6 ± 29.1 | 38.6 ± 25.4 | 0.62 | 31.4 ± 10.3 | 39.1 ± 38.6 | 0.96 | 34.5 ± 20.9 | 38.7 ± 31.7 |  |

Abbreviation: iC1e/K, inhibitor of C1 esterase/kallikrein

**Supplementary Table 6.** Adverse effects during intervention.

| **Adverse Effect** | **Total** | **Group 1** | **Group 2** | **Group 3** | **P** |
| --- | --- | --- | --- | --- | --- |
| Increased ASLT/ALT, n (%) | 14 (46.6) | 4 (40) | 5 (50) | 5 (50) | 0.88 |
| Diarrhea n (%) | 5 (16.6) | 1 (10) | 2 (20) | 2 (20) | -- |
| Nausea, n (%) | 3 (10) | 1 (10) | 0 (0) | 2 (20) | -- |
| Vomits, n (%) | 3 (10) | 1 (10) | 1 (10) | 1 (10) | -- |
| Hyperbilirubinemia, n (%) | 3 (10) | 2 (20) | 1 (10) | 0 (0) | -- |
| Bradycardia, n (%) | 2 (6.6) | 1 (10) | 0 (0) | 1 (10) | -- |
| Arrhythmia, n (%) | 0 (0) | 0 (0) | 0 (0) | 0 (0) | -- |

**Supplementary Figure 1.**
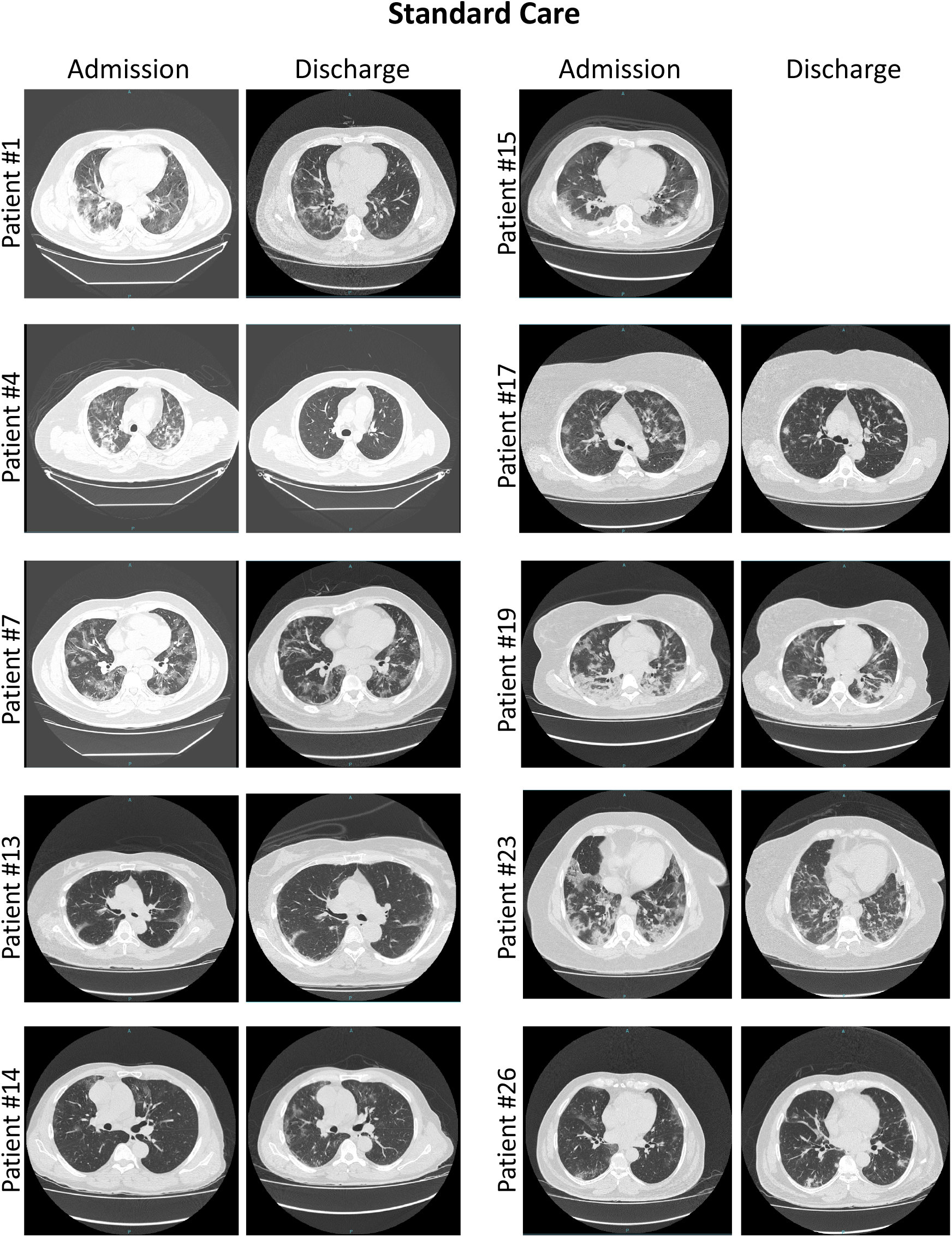
Computed tomography lung scans obtained from patients randomized to standard care. Representative images were selected from the most affected region of the lungs. Scans were obtained at admission and discharge. Patient identification refers to randomization sequence as defined in Supplementary Table 2. Patient #15 missing discharge image is due to death.

**Supplementary Figure 2.**
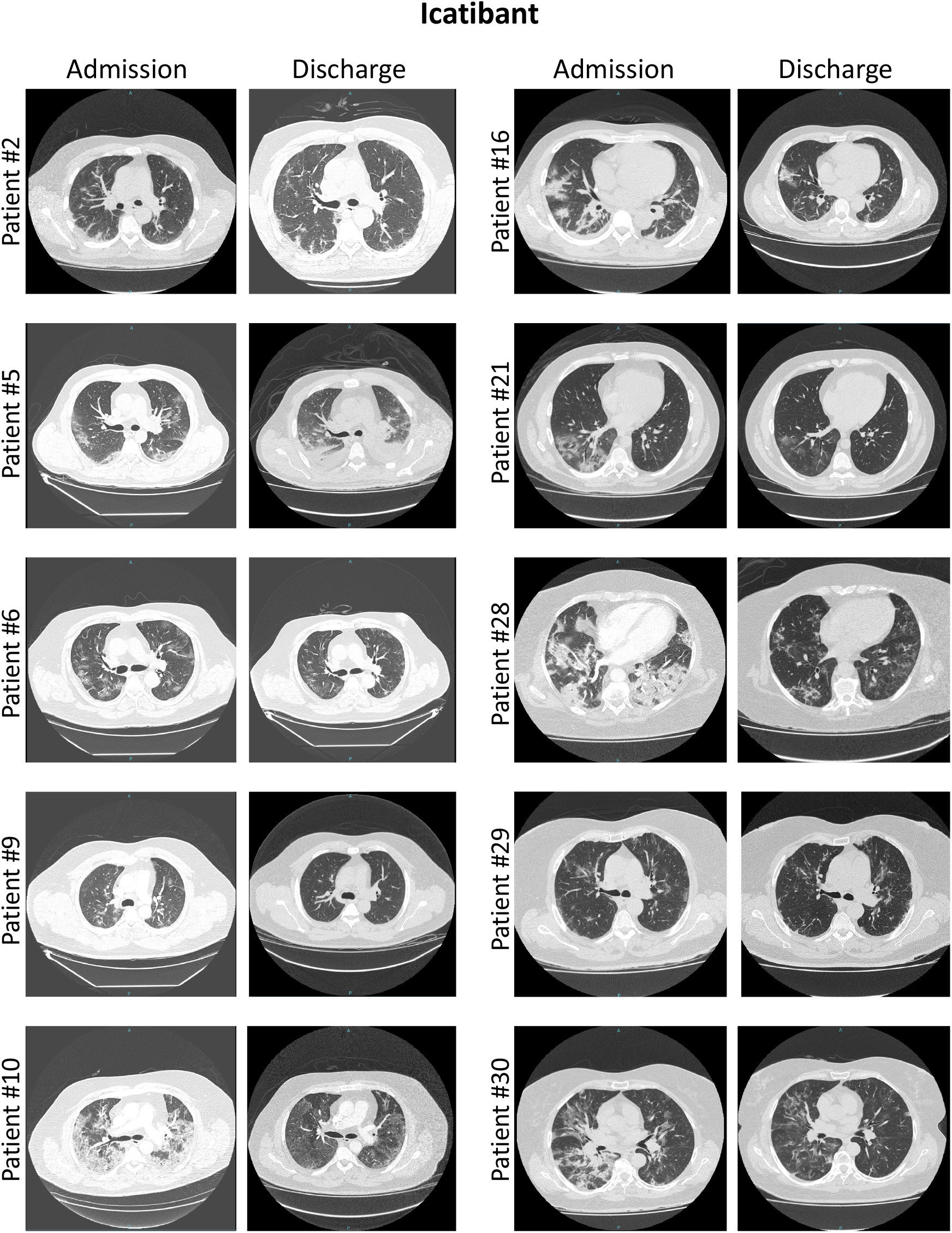
Computed tomography lung scans obtained from patients randomized to icatibant. Representative images were selected from the most affected region of the lungs. Scans were obtained at admission and discharge. Patient identification refers to randomization sequence as defined in Supplementary Table 2.

**Supplementary Figure 3.**
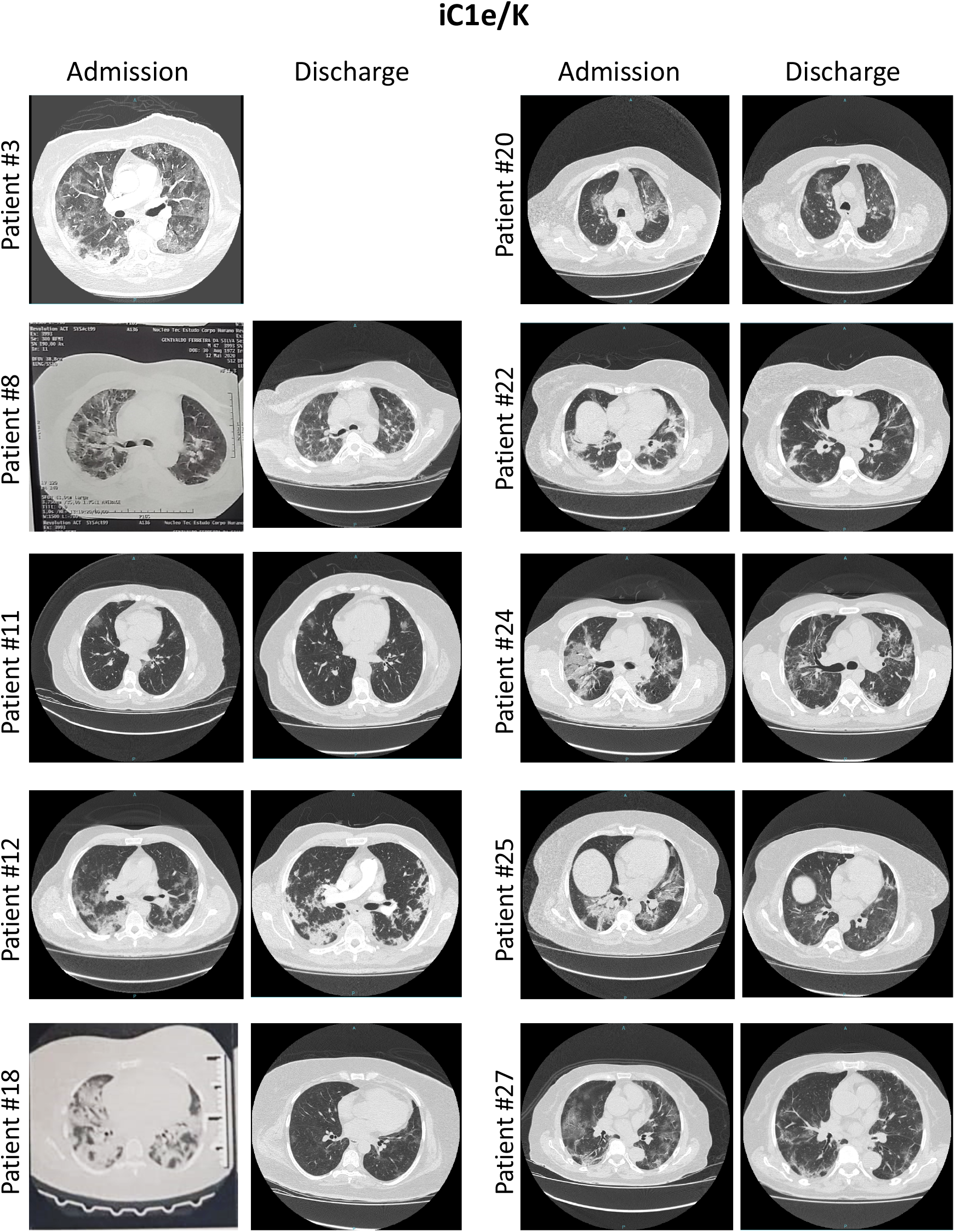
Computed tomography lung scans obtained from patients randomized to inhibitor of C1 esterase/kallikrein (iC1e/K). Representative images were selected from the most affected region of the lungs. Scans were obtained at admission and discharge. Patient identification refers to randomization sequence as defined in Supplementary Table 2. Patient #3 missing discharge image is due to death.

